# Auto tuning SIR model parameters using genetic algorithm

**DOI:** 10.1101/2021.07.19.21260722

**Authors:** L. Kristen, Fei Liu

## Abstract

Earlier studies [1] comparing Covid-19 simulations using extended SIR model [2] with observed new cases in New Jersey and United States showed good agreement between simulated results and observational data. The parameters of the SIR model controlling the behavior of the model have to be manually adjusted until the modelled results and observations reach good agreement. The parameter tuning process is tedious and time consuming. In this work, we have developed an approach using genetic algorithm [3] to automatically select the most optimal set of parameters to minimize the residual between simulated result and observational data. The parameter tuning process applying SIR model can now be automated without tedious and time consuming manual intervention.

## 1 Introduction

In December of 2019, Covid-19 began to spread rapidly in China, and would eventually reach 24 countries in just less than two months. It’s symptoms range from headaches and fatigue to difficulty breathing and coughing. Due to these symptoms also being symptoms of a fever or a cold, it is often difficult to tell whether or not a person has Covid-19 prior to taking the right precautions, such as quarantining. Covid-19 may not even present itself with any symptoms at all. Covid-19 spreads from person to person through aerosols in the air, anyone within 6 feet of an infected person can breathe in aerosols carrying the virus and become infected. It’s currently estimated that the death rate of Covid-19 is about 3%. For these reasons, Covid-19 has caused a worldwide pandemic. Luckily, vaccines and treatments have been developed to control the spread of the virus through studying the patients that have recovered as well as patients who have passed away due to Covid-19. However, even with the new vaccines, Covid-19 is still a danger to the world as new strains of the virus are evolving, as shown in India currently. In order to take preventative actions against Covid-19, people began to close public transportation and schools in order to reduce the amount of large population gatherings. It is clear to see that areas that did not take precautions to the virus are facing the consequences as hospitals began to lose its supply of resources such as ventilators, testing kits, and more.

Medical professionals are able to make more efficient and beneficial decisions in such situations mentioned previously through mathematical modeling. SIR model is such a mathematical model that can be used to assess how quickly the Covid-19 virus can spread in a population and to evaluate the strength of the Covid-19 infection.

One interesting application of SIR simulation is to examine the effect of control measure on the infected new cases in a population. Control measures can reduce the size of susceptible population and slow down or even eliminate the increase of new cases. control measures include wearing masks, observing social distancing, vaccination, etc. Having a good understanding how control measure can affect the infection rate in a population has both theoretical and practical value.

The observed new cases data from New Jersey and US have been used to set up SIR model simulation. It can be seen [1] the New Jersey and US new cases show interesting patterns that can be potentially explained by quarantine effect. The result indicates control measure in New Jersey and the US can explain the variability in the observed daily new cases.

In order to fit the observed new cases and model simulation, the traditional SIR model has been extended to the following equations:

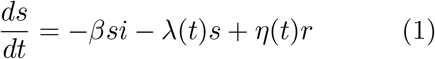

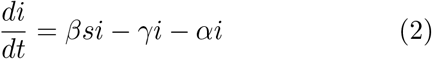

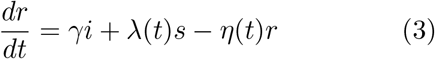

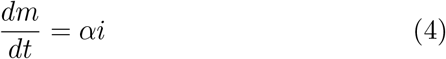

The *s, i, r* variables are the traditional susceptible, infected, and recovered population size. In the extended SIR model, a new category of pollution *m* represents mortality and *α* is the mortality rate of the virus. The effect of time dependent control measure is modelled through the function *λ*(*t*). The equations are solved simultaneously by integrating each variable in the same course of observational period from March 2020 to September 2020.

The ability of the the user to implement new parameters into the SIR model provides greater flexibility in producing more accurate results. However, in previous work given that these parameters must be manually set up to the most optimal conditions, it causes the process of identifying these parameters to be time consuming. For example, when calculating the simulated new case numbers compared to the actual observed new case numbers, the parameters for the simulated new case were found through trial and error until both the simulated and observed new cases were similar to one another. Where as with the SIR model, these parameters could be easily calculated and adjusted through automated genetic algorithm.

## 2 Genetic Algorithm

Humans are constantly attempting to learn more about the world and prepare for what is to come and genetic algorithm (GA), as part of Machine Learning techniques, can achieve this goal. GA [3], [4] is used to predict the evolution of a specific problem through evolutionary programming. Some examples of GA being implemented in the real world includes designing architecture, building automobiles, organizing traffic, etc. In order to find optimal solutions to these problems, different solutions based on slightly different conditions can be evaluated through GA.

In evaluating the success rate of chemotherapies for patients, GA allows established treatments [6] to be altered according to specific patient rather than beginning a new treatment. Alterations that can maximize an objective function based on the treatments are chosen through GA.

GA has enhanced the performance of Information Retrieval System (IRS), allowing users to attain items of information faster than other strategies such as cosine similarity fitness function. [7] According to each user’s queries, specific pieces of information are suggested through GA’s automatic similarity measure learning and matching function, thus producing more sophisticated suggestions.

Enhancing productivity and visualization, GA optimizes the problems of engineering design. [8] GA allows designers to evaluate design variables without constraints as well as to reduce the cycle of designing visual models through dynamical graphics.

In managing health care in hospitals regarding patient servicing, satisfaction, and costs, GA provides greater efficiency by shortening the waiting list, lowering the vacancy rate, and increasing the number of patients discharged. [9] The most optimal staff rotation schedule is chosen through GA to achieve these results.

In the same study by [9], leveraging GA’s predictive technology, they are able to predict the outcomes without first performing surgery. GA gives them greater insight whether to deny the surgery or to weigh the risks when they discuss the surgery options with the patients.

The concept of GA originates from how biological genes evolve in nature. In the process of evolution, genes can mutate and produce generations with slight differences in their genetic coding. These generations can be considered as different solutions that compete and optimal solutions can emerge as winners of evolution.

GA follows the same pattern and uses genetic encoding composed of necessary parameters of the problem in an automated approach to determine optimal solution in specific conditions, which otherwise would be too difficult to determine manually. [5] GA finds the fitness, known as the residual, of a given set of genes. The fitness of a gene is based on how well the gene works as a solution and is measured on how quickly it is able to find a solution, how well the gene uses it’s resources, and more. With these results, GA analyzes what generation of genes will be produced next. The greater the fitness score, the greater the likelihood of the gene being chosen for selection in recombination. In order to form the next generation, the next step in GA would be recombination. Recombination includes the recombining of genes to represent reproduction in the real world. However, while genetic material is being combined during crossover, mutations also occur in genes which are random. GA continues this process, producing numerous generations, until the genes in that population provide a solution to the given conditions.

## 3 Model Setup

In the previous work of validating the choice of the parameters, the modeler has to visually examine the simulated new cases and repeatedly compare with observation until a satisfactory set of parameters is finally chosen. Such repetitive task of pattern recognition is well suited for machine learning algorithms such as GA. In this study we apply GA to automate parameter selection process so that the time consuming visual validation work can be handled by computers.

To facilitate GA, we have chosen three parameters to be muted as the model evolves. The three parameters are *β, λ*, and *γ* used in the SIR model. Even though slight change to these parameters can significantly change the outcome of the model simulation, GA is able to continuously reduce the difference between simulated and observed new case results by choosing the better set of parameters between two successive generations.

The model starts with an relatively arbitrary set of parameters, during each generation, the genetic parameters are randomly muted. In the simplest case, the parameters either increase or decrease by *δ* = 0.1% randomly. The SIR model then computes predicted new cases using the muted parameters. It has been found that the convergence of the algorithm is sensitive to the perturbation of the parameters, e.g. altering the parameters by up to 1% can lead to divergence of the result.

In order to evaluate the quality of the mutated parameters between successive generations, a target function then calculates the accumulated residuals between simulated result and observed result.

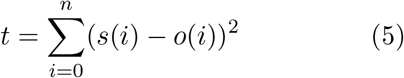

Where *i* is the number of days for the SIR model simulation, *s*(*i*) is the number of simulated infected new cases and *o*(*i*) is the number of observed infected new cases. The target function *t* calculates the accumulated residual between simulated and observed new cases for the *n* days where both observations and simulation results are available. To improve the model simulation result, we choose the set of genetic parameters that reduces the target function.

SIR model parameters are mutated during each generation through random chance, a random number between 0 and 1 is generated, depending on the magnitude of the random number, for example *β* can be adjusted by

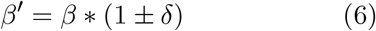

where *δ* measures the magnitude of mutation. Because the residual tends to plateau after a few hundreds of generations, a time independent magnitude of mutation *δ* can lead to oscillatory behavior (Figure 1). To reduce the oscillatory behavior when the residual plateaus, the magnitude of mutation is reduced by

**Figure 1:**
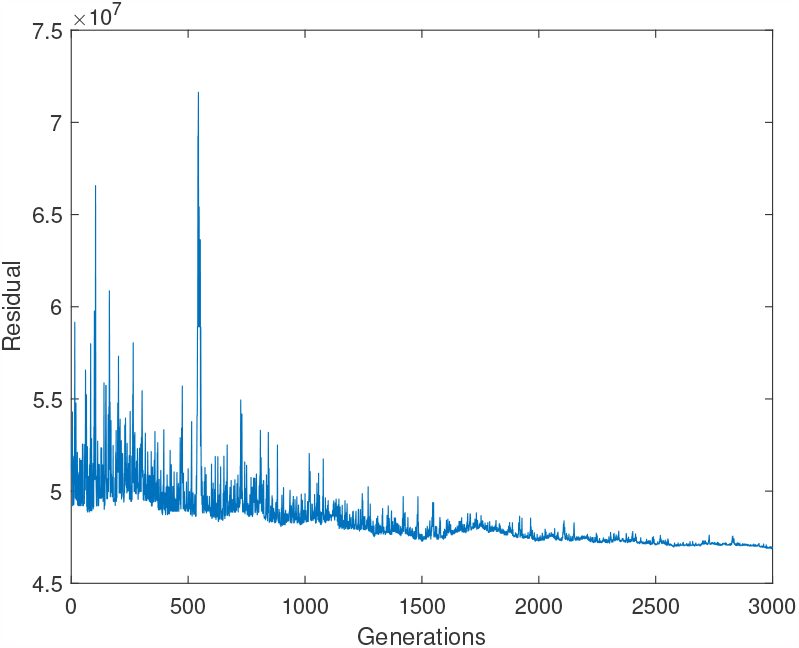
The evolution of residual starting from *β* = 0.28, *γ* = 0.07, *λ* = 0.026. After 6000 evolutions, the residual is reduced to 4.93e7 with optimized parameters at *β* = 0.2487, *γ* = 0.0431, *λ* = 0.0283

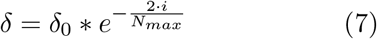

where *i* represents the i-th generation during application of genetic algorithm and *N*_*max*_ represents the maximum number of iterations chosen for the simulation. The exponential decay function ensures the genetic mutation during successive generations become smaller over time and allows the algorithm to minimize the residual.

## 4 Result

We first start with the parameters hand tuned and apply GA to further optimize the match between the observed and simulated new cases in New Jersey. Figure 1 shows the residual calculated using (Equation 5). Due to the exponential decay of the mutation magnitude *δ*, the residual first oscillates rapidly but stabilizes as the number of generation increases.

It should be noted that due to the probabilistic nature of the genetic algorithm, the residuals are not reproducible between simulations. At the same time, since the number of generations simulated is large, the result approaches the same behavior statistically (large number theorem).

We also experimented adding the ’shift’ parameter as part of genetic mutation. The result is shown in (figure 5). The shift parameter is the day at which the control measures take effect [1]. It can be seen the shape of the simulated curve is very similar to observed cases as seen in figure 3.

**Figure 2:**
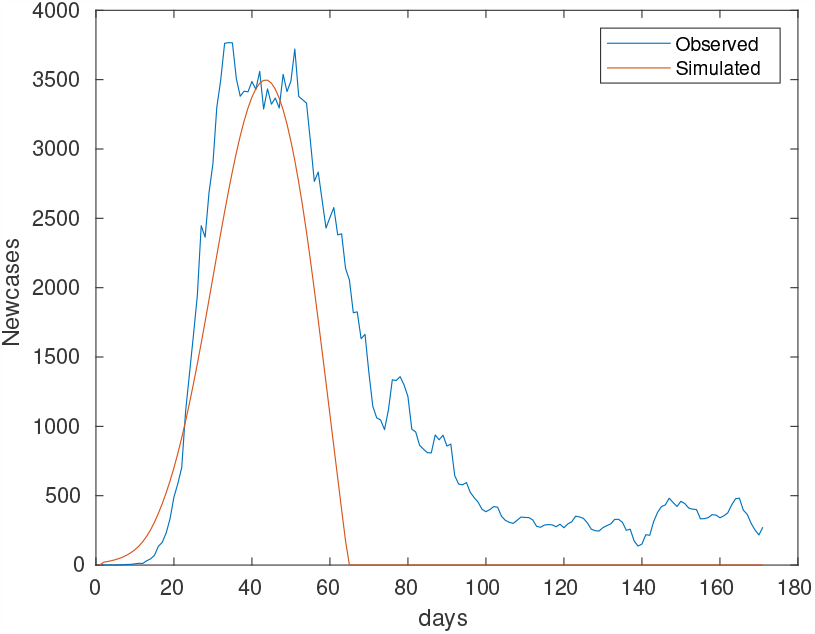
Observed and Simulated new cases using the parameters *β* = 0.28, *γ* = 0.07, *λ* = 0.026 as in [1].

**Figure 3:**
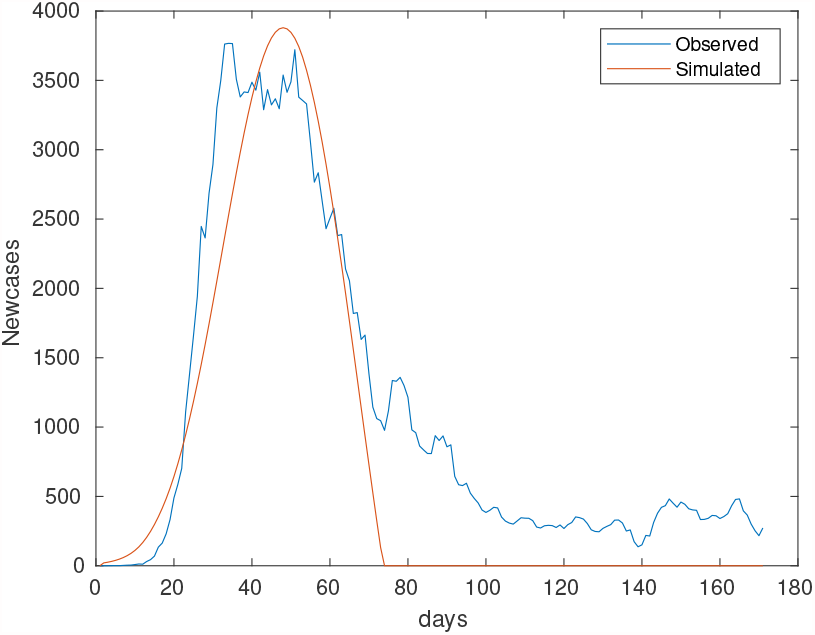
The evolution of residual starting from *β* = 0.28, *γ* = 0.07, *λ* = 0.026. After generating the residual twice, each with 3000 generations, resulting in a total of 6000 generations, the residual is reduced to 4.93e7 with optimized parameters at *β* = 0.2487, *γ* = 0.0431, *λ* = 0.0283

**Figure 4:**
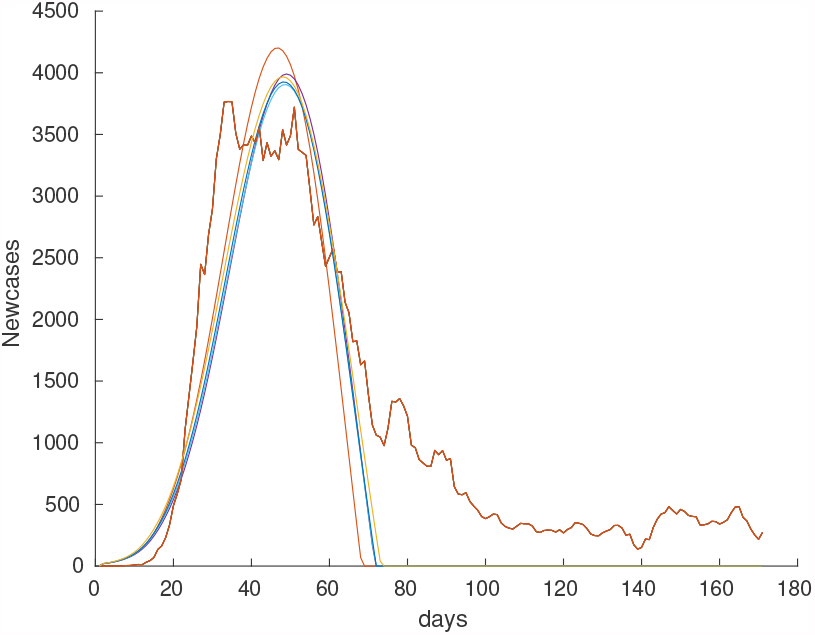
The evolution of residual starting from *β* = 0.35, *γ* = 0.09, *λ* = 0.036, shift = 15. After 6000 generations, the residual is reduced to 4.4983e7 with optimized parameters at *β* = 0.2602, *γ* = 0.0491, *λ* = 0.026, shift = 13.01

**Figure 5:**
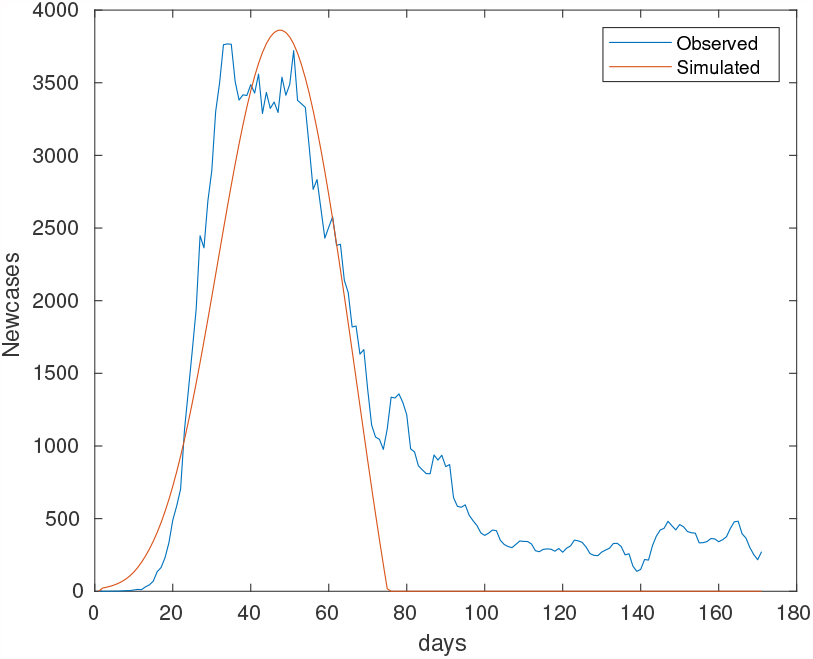
The evolution of residual starting from *β* = 0.28, *γ* = 0.07, *λ* = 0.026, *shift* = 15. After 6000 evolutions, the residual is reduced to 4.4983e7 with optimized parameters at *β* = 0.2595, *γ* = 0.0361, *λ* = 0.0292, *shift* = 11.2210

It can be seen from (figure 4), the shape of the simulated new cases curve changes as the number of generation increases with the peak approaching the observed new cases. The green curve is after 6000 generations.

The algorithm is able to reduce the residual and compute a better set of parameters to match the observed and simulated cases through iterations. For example, we can start the parameters as *β* = 0.35, *γ* = 0.09, and *λ* = 0.036. Then we successively run the algorithm first with 1000 generations 3 times and finally with 3000 generations once. The result is shown in (figure 4). Both figure 3 and 5 show better fit than the original parameters used in figure 2.

**Table 1:** Residual using varying initial values of *β, γ, λ*, the algorithm can minimize the Residual regardless of the initial values used.

| $\beta$ | $\gamma$ | $\lambda$ | Residual |
| --- | --- | --- | --- |
| 0.28 | 0.07 | 0.026 | 5.10E+07 |
| 0.29 | 0.08 | 0.036 | 5.30E+07 |
| 0.3 | 0.09 | 0.046 | 5.47E+07 |
| 0.31 | 0.10 | 0.056 | 5.68E+07 |
| 0.32 | 0.11 | 0.066 | 5.33E+07 |
| 0.33 | 0.12 | 0.076 | 5.61E+07 |
| 0.34 | 0.13 | 0.086 | 5.81E+07 |
| 0.35 | 0.14 | 0.096 | 5.55E+07 |
| 0.36 | 0.15 | 0.106 | 6.09E+07 |

## 5 Conclusion

To reduce the amount of manual intervention needed to tune the SIR model to fit observed new cases, we set up SIR model to use mutated parameters each generation. Those mutations that lead to reduced residual are kept as new basis to perform subsequent mutation and simulation.

It’s been found with constant magnitude of mutation, the residual between observed and simulated new cases show oscillatory behavior. To reduce the oscillation, the magnitude of mutation is allowed to decay over generations, reducing the oscillation in the optimization process. This is analogous to finding the minimum of a terrain, where we first run with great strides to a low land, once we arrive at the low land, we travel slowly to find the minimum at the low land. This exponential decay behavior has significantly improved the stability of the algorithm while being able to reduce the residual by exploring large variations of the parameter space initially.

The mutations of the information generally leads to reduced residual regardless of the initial parameters fed into the SIR model. For the specific case, the residual can be reduced to about 5e7 regardless of initial conditions quickly. Subsequent reduction of the residual becomes slower and requires more generations to be more effective.

Using the original numbers from [1], we can see the genetic algorithm is able to easily further optimize the parameters used to better match simulation with observation in a matter of minutes where the original set of parameters took days of running the code and comparing with observation to unveil.

The efficiency of the genetic algorithm at discovering optimal parameters in model simulation is demonstrated by its ability to match the simulated result with observation with minimal human intervention. Such approach can be applied to any activity where large number of trials and errors are needed to match patterns and optimize functions heuristically.

## Data Availability

All data used in the study are in public domain and can be accessed at https://github.com/CSSEGISandData/COVID-19

